# Diagnostic Potential for IgM Antibody Detection by the DPP Syphilis TnT Assay in Neonates at Risk for Congenital Syphilis

**DOI:** 10.64898/2026.01.05.26343454

**Authors:** Irene A. Stafford, Lierni Ugartemendia Ugalde, Laura M. Goetzl, Analuisa Mosqueda, Sabrina DaCosta, Dhammika Gunasekera, Mark Rivieccio, Javan Esfandiari, Konstantin P. Lyashchenko

## Abstract

**Background:** Neonatal IgM antibodies reflect an in-utero immune response to *Treponema pallidum* and may offer added diagnostic value. This study evaluated the test performance of treponemal IgM levels measured by the research-use-only (RUO) DPP Syphilis TnT point-of-care (POC) assay for CS risk stratification.

**Methods:** Conducted from May 2023 to May 2025, this study tested neonatal serum samples from infants born to mothers with syphilis using the DPP Syphilis TnT RUO POC assay, which reports treponemal and nontreponemal IgM levels as relative light units (RLU). Neonates were classified as *Confirmed Proven* or *Highly Probable CS*, *Possible CS*, or *CS Less Likely* per guidelines; 23 neonates without maternal syphilis served as controls. Treponemal IgM levels were compared across categories using nonparametric tests and ordinal logistic regression. Diagnostic performance used prespecified cutoffs, with agreement assessed against neonatal RPR.

**Results:** Twenty-two maternal–neonatal dyads were included. Mean treponemal IgM levels rose with CS severity, peaking in the high-risk group (*Possible* or *Confirmed Proven/Highly Probable* CS: 29.9 ± 20.6 RLU) versus *CS Less Likely* (17.5 ± 20.8 RLU) and controls (3.5 ± 0.8 RLU; p<0.05). Higher IgM levels independently linked to elevated CS risk (OR 1.10 per 1 RLU; p=0.0025). At ≥10 RLU cutoff, treponemal IgM detected 88.9% of high-risk neonates, with 76% agreement to neonatal RPR.

**Conclusion:** The DPP Syphilis TnT RUO POC assay’s treponemal IgM levels discriminated CS risk categories effectively and may supplement current algorithms to improve neonatal CS stratification.

## Introduction

According to 2022 World Health Organization (WHO) global surveillance data, the estimated number of congenital syphilis (CS) cases is increasing, with over 700,000 cases estimated worldwide, exceeding the WHO global mother to child transmission (MTCT) threshold by over 10-fold (1). The adverse birth outcomes related to maternal and CS are profound, with over 390,000 adverse birth outcomes reported in 2022, including 150,000 fetal deaths resulting from untreated maternal infections worldwide (1–4). For infected infants born alive, the estimated cost of neonatal intensive care for one newborn with CS in the United States exceeds $55,000, excluding quality of life or activities-of-daily-living expenses for affected families (5). Although guidance for adult infection is well-established and evidence-based (6,7), the current diagnostic strategies for neonatal infection rely on a risk-based algorithm dependent on maternal diagnosis and treatment history, exam findings, placental evaluation, and comparative levels of maternal and neonatal nontreponemal (NT) antibodies detected by Venereal Disease Research Laboratory (VDRL) or rapid plasma reagin (RPR) tests at delivery (6,7). All these individual criteria lack strong test performance, and when taken together, are used to estimate scenario-based CS risk categories to inform treatment strategies (e.g., *1) Confirmed Proven or Highly Probable CS, 2) Possible CS, 3) CS Less Likely, and 4) CS Unlikely*), according to the current Centers for Disease Control and Prevention (CDC) STI Treatment Guidelines (6,7). Results of RPR testing are often challenging to interpret due to maternal IgG antibody interference, therefore necessitating between 2 – 15 months of lengthy follow-up with RPR monitoring to ensure clearance of maternal IgG antibody and absence of congenital infection (6–8). The NT serologic assays require venipuncture, specialized equipment, and off-site moderately complex laboratory settings, which often lead to incomplete care cycles with poor follow-up for results management or treatment (6,7). In contrast, point-of-care (POC) tests like the FDA-cleared Syphilis Health Check or Dual Path Platform (DPP) HIV-Syphilis Assay enable decentralized testing in clinics, emergency departments, shelters, and other non-clinical environments by trained staff (9–11). These are considered first-line options for adult syphilis testing in resource-limited global settings, given the advantage of a 15–20-minute turnaround time from testing to results, which can facilitate same-day treatment for early infection, prevent MTCT and potentially cure fetal disease when used in antenatal screening (9–11).

It has been demonstrated that IgM antibodies detectable in neonates at risk for CS are indicative of a fetal immune response to active *Treponema pallidum* infection *in utero* (12). While IgM-specific assays for CS have demonstrated sensitivities of 83–100% in symptomatic cases and specificities up to 100%, their overall performance varies (12–15). This reported variability of diagnostic accuracy can be explained by anti-treponemal testing performed prior to seroconversion in cases of early CS, or after the clearance of antibodies following treatment for early-stage fetal infection. Additionally, there is no gold-standard comparator or FDA-cleared anti-treponemal IgM test available in the US (12–15). Given these limitations, the use of a treponemal IgM test for CS diagnosis has not been adopted in the US or recommended by the CDC or WHO (1,7). Further validation is needed for POC applications in neonates to account for maternal IgG interference to potentially optimize clinical algorithms using a rapid treponemal IgM test.

The objective of this pilot study was to assess the diagnostic value of IgM responses detected by the Chembio DPP Syphilis TnT research-use-only (RUO) point-of-care (POC) Assay in neonates categorized at birth into various CS risk scenarios, per the 2021 CDC STI Treatment Guidelines (7). The neonatal IgM antibody levels and reactivity rates across the risk groups were also analyzed in comparison with the newborn RPR test results used routinely for CS diagnosis.

## Materials and Methods

The study was conducted at the University of Texas Health Science Center-obstetric tertiary care center in Houston, Texas, from May 2023 to May 2025. Pregnant individuals aged 14-45 with a confirmed live intrauterine pregnancy at greater than 16 weeks gestational age (GA), presenting for routine prenatal care and with a diagnosis of syphilis during the index pregnancy were eligible for enrollment. The diagnosis of syphilis was made using the laboratory-based reverse syphilis testing algorithm, which involves a treponemal test, such as an enzyme immunoassay (EIA), followed by RPR test for confirmation of active infection. A secondary treponemal test, T. pallidum particle agglutination (TP-PA), was used when the reverse algorithm produced discrepant results (7). All patients underwent a detailed health history and physical exam for the staging of syphilis and were treated with benzathine penicillin G according to stage (*primary, secondary, early latent, late latent*) per the CDC STI Treatment Guidelines (7). Infection status was confirmed with the regional health department. Exclusion criteria included pregnancies with syphilis treated prior to the pregnancy, or if pregnancy was complicated by a fetal demise (7). All consented participants underwent repeat testing at 28 weeks and delivery according to the Texas Health and Safety Code 81.090 unless more frequent testing was warranted. All pregnancies were managed according to the standard of care, regardless of participation in the study. All maternal and neonatal infection related data were recorded.

The study population consisted of 45 newborns including 22 neonates at risk for CS (born to mothers diagnosed with syphilis during pregnancy) and 23 no-risk neonates (no exposure to maternal syphilis) used a negative control group. Within 48 hours of delivery, 50 -75 microliters (μL) of neonatal serum were collected and stored frozen at -70⁰ C until testing by the DPP Syphilis TnT RUO POC Assay (Chembio Diagnostics, Inc., Medford, NY) and Trep-Sure™ Syphilis Total Antibody EIA (Trinity Biotech, Jamestown, NY).

The Chembio DPP Syphilis TnT Assay originally developed for syphilis testing in adults is designed to have four separate test lines for differential semi-quantitative detection of IgM and IgG antibodies to treponemal and NT antigens. The treponemal test lines use the same proprietary multi-epitope recombinant protein as employed in the DPP HIV-Syphilis Assay product (PMA Approved, BP180191;10/2020), whereas the NT test lines use the CDC-licensed VDRL antigen with in-house improved liposomal formulation. To run the assay, 10 uL of serum was mixed with 5 drops (150 uL) of DPP Syphilis TnT Running Buffer in the test kit provided sample vial. A 125 uL portion of this sample mixture was transferred to test device Well #1 using the transfer pipette provided in the test kit. After waiting 5 minutes, 10 drops (300uL) of DPP Syphilis TnT Running Buffer were added to test device Well #2. Results were interpreted using the Chembio DPP Micro Reader II at 15 minutes after the Running Buffer was added. The DPP Micro Reader II was configured to report a qualitative result for neonatal IgM results interpreted as reactive or non-reactive based on pre-established cutoff values of 10 relative light units (RLU) for each test antigen and the level for each antibody class (IgM and IgG) for treponemal and NT antibodies as independent outputs, expressed as RLU. All results were recorded for analysis.

At birth, neonatal providers assigned the clinical CS risk scenario following the CDC Guidelines and American Academy of Pediatrics standards (7,18) prior to experimental testing. All aliquots were run in duplicates, and the investigators were blinded to the neonatal CS risk category assigned at birth. The protocol was approved by the Institutional Review Board of the McGovern Medical School (HSC-MS-25-0170). All participants provided written informed consent to participate in the research study.

Descriptive statistics were used to summarize participant data comparing all at-risk categories (*Confirmed Proven or Highly Probable CS, Possible CS, and CS Less Likely*) and the negative controls. For data analysis, the *Possible CS and Confirmed Proven or Highly Probable CS* results were combined in one group given their relatively low numbers and high-risk status (7). Summary statistics (median and range) were calculated for maternal and neonatal RPR test results. Kruskal–Wallis tests were used to evaluate differences in treponemal and NT mean IgM levels detected by the Chembio DPP Syphilis TnT Assay across CS categories. Ordinal regression was performed to determine relationship between treponemal IgM values and CS risk category. Pairwise Mann–Whitney U tests with Bonferroni correction were performed for post-hoc contrasts. Receiver operating characteristic (ROC) analysis was used to evaluate the diagnostic performance of treponemal and NT IgM individually. A composite IgM predictor was constructed using multivariable logistic regression incorporating both, and ROC analysis of the resulting score was performed to assess incremental discrimination. Diagnostic performance was assessed using 2×2 contingency tables, with sensitivity and specificity calculated for treponemal IgM cutoffs. Given the lack of a diagnostic gold-standard test for CS, the clinical CDC diagnostic criteria were used as the comparator for analysis and the *Possible or Confirmed Proven or Highly Probable CS* group was considered test-positive as described above. Exact 95% confidence intervals were estimated using the Wilson method. Agreement between neonatal treponemal IgM and RPR test results at delivery was evaluated using categorical agreement analysis. Overall percent agreement was calculated, and agreement beyond chance was quantified using Cohen’s kappa (κ All statistical analysis was performed using STATA software, version 17 (College Station, TX).

## Results

From May 2023 to May 2025, a total of 22 maternal–neonatal dyads met inclusion criteria and had complete clinical data along with available serum for both experimental assays. These at-risk neonates were classified as *Confirmed Proven or Highly Probable CS (n=6), Possible CS* (n=3) and *CS Less Likely* (n=13). Table 1 demonstrates baseline infection related characteristics of the enrolled maternal-neonatal dyads. Most of the infected women were diagnosed and treated for syphilis between 16 – 26 weeks gestational age (n=14, 64%) and delivered at term (n=17, 77%). Twenty-three neonates without exposure to maternal syphilis were used as negative controls.

**Table 1.** Maternal and Neonatal Syphilis Characteristics and RPR Titers at Delivery Maternal Information.

| Maternal Syphilis Stage | N | Median NT Titer | RPR Titer Range |
| --- | --- | --- | --- |
| Early Syphilis | 3 | 16 | 4–128 |
| Late / Late-Latent | 19 | 2 | NR–32 |

**Neonatal Information** DPP TnT IgM Assay for Congenital Syphilis
| Neonatal CS Category | N | Median NT Titer | RPR Titer Range |
| --- | --- | --- | --- |
| *CS Less Likely* | 13 | 1 | NR–4 |
| *Possible CS* | 3 | 2 | NR–4 |
| *Confirmed Proven/ Highly Probable CS* | 6 | 8 | NR–16 |

Neonatal treponemal IgM antibodies were detected by the Chembio DPP Syphilis TnT Assay in all at-risk categories, with the highest mean levels observed in the high-risk CS group (*Possible CS* and *Confirmed Proven or Highly Probable CS* combined) (29.9 ± 20.6) compared to the *CS Less Likely* cohort (17.5 ± 20.8) or the negative controls (3.5 ± 0.8) (p < 0.05) (Table 2). NT IgM also differed significantly across CS categories but with less pronounced differences. Figure 1 displays treponemal and NT IgM values according to CS category.

**Figure 1.**
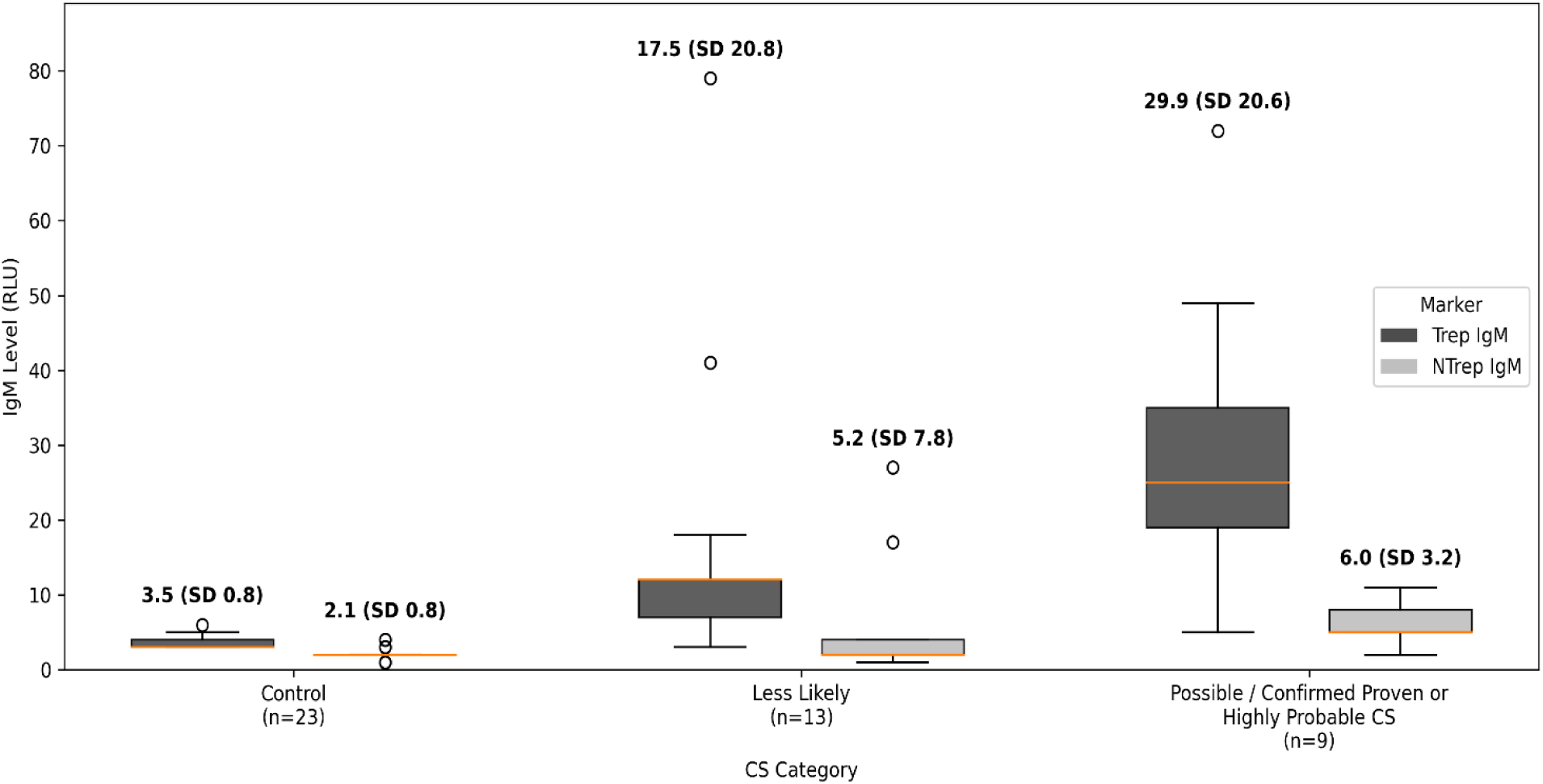
Box plots display neonatal treponemal and nontreponemal IgM levels, expressed as relative light units (RLU), stratified by congenital syphilis (CS) category (Control, *CS Less Likely,* and *Possible / Confirmed/ Highly Probable CS).* The *Possible* and *Highly Probable/Confirmed CS* categories were combined for analysis. Boxes represent the interquartile range (IQR), center lines indicate medians, and whiskers extend to 1.5× the IQR. Mean values with standard deviations (SD) are shown above each category. Differences across categories were assessed using the Kruskal–Wallis test.

**Table 2.** Treponemal and Nontreponemal IgM Levels by Congenital Syphilis Category *(Possible and Confirmed Proven or Highly Probable CS* combined)

| CS Category | n | Treponemal<br>IgM Mean<br>(SD) | Treponemal<br>IgM Median | Treponemal<br>IgM Min–Max | NT IgM<br>Mean<br>(SD) | NT IgM<br>Median | NT IgM<br>Min–Max |
| --- | --- | --- | --- | --- | --- | --- | --- |
| Negative Control | 23 | 3.5 (0.8) | 3 | 3–6 | 2.1 (0.8) | 2 | 1–4 |
| *CS Less Likely* | 13 | 17.5 (20.8) | 12 | 3–79 | 5.2 (7.8) | 2 | 1–27 |
| *Possible/Confirmed*<br>*/Highly Probable CS* | 9 | 29.9 (20.6) | 25 | 5–72 | 6.0 (3.2) | 5 | 2–11 |

*Possible and Confirmed Proven or Highly Probable CS* categories were combined for analysis. Treponemal IgM and NT IgM levels are reported as Relative Light Units (RLU).

In ordinal logistic regression including controls, higher mean treponemal IgM levels were significantly associated with increasing CS risk category (OR 1.10 per 1 RLU, 95% CI 1.04–1.18; p=0.0025), corresponding to an OR of 1.64 per 5 RLU (95% CI 1.19–2.26).

Treponemal IgM was the primary driver of diagnostic sensitivity for CS detection. Using a cutoff of ≥ 10 RLU, the treponemal IgM positive results obtained with the Chembio DPP Syphilis TnT Assay identified 8/9 (88.9%) neonates in the high-risk groups and 8/13 (61.5%) newborns in the *CS Less Likely* scenario. Risk stratification among high-risk neonates (*Possible CS and Confirmed Proven or Highly Probable CS*) could be enhanced by applying a higher treponemal IgM cutoff of ≥19 RLU. At this threshold, test specificity increased to 94.4%, with a corresponding reduction in sensitivity to 77.8%, potentially resulting in two missed high-risk CS cases. Neonates classified as *CS Less Likely* were included in the negative group for diagnostic performance analyses; however, these infants represent an exposed population, and misclassification cannot be fully excluded.

In contrast, NT IgM demonstrated significantly lower seroreactivity rates and weaker differentiation between the risk categories. Using a cutoff ≥ 10 RLU, this biomarker detected 2/9 (22%) high-risk neonates, 2/13 (15.4%) low-risk neonates, and none among 23 no-risk neonates. The four newborns with NT IgM antibodies found were also RPR reactive (titers ranging from 1:1-1:4) and positive for treponemal IgM antibodies detected by the Chembio DPP Syphilis TnT Assay at relatively high levels (50.5 ± 11). Although a combined treponemal and NT IgM metric improved overall discrimination as assessed by ROC analysis, it did not enhance the test sensitivity beyond treponemal IgM alone.

Among the 22 at-risk neonates, 15 tested RPR reactive at birth, of which 13 (86.7%) had treponemal IgM antibodies. In contrast, out of the remaining 5 RPR non-reactive newborns, only 3 (42.9%) produced this serologic biomarker. When the treponemal IgM results were stratified by RPR titers, the IgM component of Chembio DPP Syphilis TnT Assay detected all at-risk neonates (5/5) with RPR titers ≥1:4, as compared to lower treponemal IgM reactivity rate of 80% (8/10) found in the low-titer subgroup (≤1:2). At the 10 RLU cutoff, categorical agreement between treponemal IgM positivity and neonatal NT positivity at delivery was 76%, with moderate agreement beyond chance (Cohen’s κ=0.44), indicating concordance with the RPR titers used for clinical management. All syphilis-exposed neonatal samples were positive by the Trep-Sure™ Syphilis Total EIA test, confirming the presence of maternal IgG or both isotypes of treponemal antibody in neonatal serum.

## Discussion

In this pilot study of neonates at risk for syphilis infection, treponemal IgM reactivity rates and levels measured using the Chembio DPP Syphilis TnT Assay increased across clinical CS categories. Mean RLU values were significantly higher among neonates classified as *Possible CS* and *Confirmed Proven or Highly Probable CS* compared with the negative controls (∼8.5-fold) and with those classified as *CS Less Likely* (∼1.7). This was also demonstrated in ordinal regression analysis, where each one-unit increase in RLU conveyed an increased level of CS risk. Median values and overall distributional shifts also demonstrated progressively higher treponemal IgM levels with increasing CS severity. Minor nonmonotonicity in mean values was observed, attributable to small sample sizes and greater variability within the high-risk *CS* group.

NT IgM levels also differed across CS categories, although less significantly if compared with treponemal IgM. Overall, treponemal IgM demonstrated superior discrimination across CS risk categories and aligned with laboratory-based RPR test performed at delivery, supporting its potential utility for neonatal CS risk stratification.

Two neonates had notable clinical contexts. One neonate classified as *Possible CS* was born to a mother with systemic lupus erythematosus and demonstrated a treponemal IgM response within the range observed for this category. Another neonate classified as *CS Less Likely* was exposed to antenatal corticosteroids for fetal lung maturation approximately two weeks prior to delivery and exhibited a relatively high treponemal IgM level (72 RLU). These observations suggest that maternal autoimmune disease or exposure to antenatal corticosteroids did not preclude development of treponemal IgM responses in this neonatal cohort.

A treponemal IgM cutoff of ≥10 RLU provided optimal sensitivity (89%), prioritizing detection of all at-risk neonates while minimizing missed cases. These findings are particularly relevant in clinical and public health settings, both globally and within the U.S., when maternal infection history, treatment status, or related information may be incomplete, unavailable, or unreliable at the time of delivery. In such contexts, neonatal biomarkers that enable sensitive, objective risk assessment independent of maternal history are critical for timely identification and management of CS. In contrast to lower IgM thresholds optimized for screening, a treponemal IgM value ≥19 RLU may have particular utility as a risk-stratification marker. At this threshold, specificity was high (94.4%), suggesting that neonates with IgM levels above this cutoff represent a population at especially high risk for CS. Although sensitivity is reduced, such values may support clinical decisions regarding escalation of evaluation, or prioritization for close follow-up, particularly when clinical RPR titers are equivocal.

The serologic response to *T. pallidum* begins with the development of treponemal IgM antibodies, which emerge earlier than the NT response in fetal, neonatal, and adult infection (12–15). Because IgM is produced in primary or recent infection and does not always persist in late-stage disease, it provides valuable temporal information that IgG cannot. In contrast, the RPR test used for clinical diagnosis and monitoring detects reagin antibodies of both IgG and IgM classes against lipoid (cardiolipin–lecithin–cholesterol) antigen (7,8). The RPR test reports a total antibody response and therefore cannot distinguish between immunoglobulin isotypes or differentiate maternal IgG transferred to fetus from neonatal IgM production. As a result, comparative RPR titers may substantially underestimate early neonatal infection (12). The clinical neonatal RPR results produced in the present study were uniformly of low titers across all CS categories and were essentially nondiagnostic. To meet the CDC-recommended diagnostic criteria for CS, an RPR titer in neonates must be at least fourfold higher than the maternal RPR titer at delivery (7). However, this approach has poor clinical sensitivity as most infected neonates have equivocal RPR titers (12). Most at-risk infants in the present study, including some classified as *Confirmed Proven or Highly Probable CS* by the clinical care team demonstrated no or very low RPR titers. In contrast, treponemal IgM values showed clear stratification across the CS risk groups and remained elevated even when neonatal RPR titers were low.

There are commercial IgM immunoblot and ELISA assays used in Europe (e.g., ViraMed and Euroimmun, Germany), all of which are CE-marked and therefore meet regulatory standards for in-vitro diagnostics in the European market. Published data show that these CE-marked IgM immunoblot assays (ViraMed, Euroimmun) demonstrate strong diagnostic accuracy, with reported sensitivities ranging from 90.0% to 94.4% and specificities between 97.1% and 99.2% when evaluated against composite or reference standards in infants and neonates with suspected CS (13–15,16,19,20). In contrast, the CE-marked Euroimmun IgM ELISA typically shows more modest sensitivity (≈ 60%) despite specificity approaching 100%. Historical studies, including those using the 19S IgM-FTA-ABS assay, similarly demonstrated that IgM-based methods may offer meaningful diagnostic value in CS (13–15,19,20). More recent work, including a 2025 study evaluating a different experimental lateral-flow POC IgM test, suggests that newer technologies may achieve high diagnostic performance with sensitivities in the 88–92% range and specificities of 96–98% when compared to a composite reference comparator and clinical CS categories (16,17).

Despite encouraging performance from CE-marked assays internationally and promising early US data using experimental point-of-care anti-treponemal IgM tests, no treponemal-IgM assay is currently FDA-cleared for CS diagnostics in the U.S. Consistent with this, routine IgM testing is not recommended by the CDC because of variable accuracy, limited clinical validation, and inconsistent correlation with disease stage (7). These considerations underscore the critical need for improved neonatal-specific diagnostic tools, particularly given that many infected infants are asymptomatic at birth, traditional RPR titers are often nondiagnostic, and that long-term serologic follow-up is difficult to achieve for many families (6,7). More accurate, rapidly deployable assays, such as the Chembio DPP Syphilis TnT Assay evaluated in the present study, would therefore represent a major advance for timely identification and management of CS.

## Strengths

This pilot study has several important strengths. First, it includes a well-characterized cohort of maternal–neonatal dyads with confirmed syphilis infection, with maternal staging performed according to current CDC STI Treatment Guidelines by clinicians and nurses with established expertise in syphilis management. Second, all neonatal specimens were collected within 48 hours of delivery and processed within one hour, ensuring high sample integrity. Third, the Chembio DPP Syphilis TnT Assay was performed by research staff who were blinded to neonatal CS categories, which were independently assigned by pediatric clinicians with expertise in congenital infections.

## Weaknesses

This study has limitations. The sample size was modest (n=21 neonates and 23 controls) and the study period relatively brief, limiting generalizability. However, the consistency of findings, statistically significant differences across CS categories, and rigorous methodology provide confidence in the internal validity of the results. Given the ongoing syphilis epidemic and the urgent clinical need for improved CS diagnostics, dissemination of these preliminary findings remains both relevant and necessary to guide further research and test development (1,7,8).

## Conclusion

Our findings demonstrate that detection of treponemal IgM antibodies by the Chembio DPP Syphilis TnT Assay in neonates may have an added diagnostic value for CS risk assessment. The potential integration of a rapid POC treponemal IgM test into the neonatal CS diagnostic algorithm would be paradigm shifting for families and clinicians, particularly given the longstanding absence of a reliable gold-standard diagnostic test for an infection associated with significant neonatal morbidity and mortality.

## Data Availability

All data produced in the present study are available upon reasonable request to the authors

## Acknowledgments

This work was supported, in part, by the Bill & Melinda Gates Foundation, Grant Number INV-INV-079080. Under the grant conditions of the Foundation, a Creative Commons Attribution 4.0 Generic License has already been assigned to the Author Accepted Manuscript version that might arise from this submission.

Deidentified data can be provided with written request and approval by all authors.

